# Species distribution modeling for disease ecology: a multi-scale case study for schistosomiasis host snails in Brazil

**DOI:** 10.1101/2023.07.10.23292488

**Authors:** Alyson L. Singleton, Caroline K. Glidden, Andrew J. Chamberlin, Roseli Tuan, Raquel G. S. Palasio, Adriano Pinter, Roberta L. Caldeira, Cristiane L. F. Mendonça, Omar S. Carvalho, Miguel V. Monteiro, Tejas S. Athni, Susanne H. Sokolow, Erin A. Mordecai, Guilio A. De Leo

**Author notes:** Corresponding author (AS).

## Abstract

Species distribution models (SDMs) are increasingly popular tools for profiling disease risk in ecology, particularly for infectious diseases of public health importance that include an obligate non-human host in their transmission cycle. SDMs can create high-resolution maps of host distribution across geographical scales, reflecting baseline risk of disease. However, as SDM computational methods have rapidly expanded, there are many outstanding methodological questions. Here we address key questions about SDM application, using schistosomiasis risk in Brazil as a case study. Schistosomiasis—a debilitating parasitic disease of poverty affecting over 200 million people across Africa, Asia, and South America—is transmitted to humans through contact with the free-living infectious stage of *Schistosoma* spp. parasites released from freshwater snails, the parasite’s obligate intermediate hosts. In this study, we compared snail SDM performance across machine learning (ML) approaches (MaxEnt, Random Forest, and Boosted Regression Trees), geographic extents (national, regional, and state), types of presence data (expert-collected and publicly-available), and snail species (*Biomphalaria glabrata*, *B. tenagophila* and *B. straminea*). We used high-resolution (1km) climate, hydrology, land-use/land-cover (LULC), and soil property data to describe the snails’ ecological niche and evaluated models on multiple criteria. Although all ML approaches produced comparable spatially cross-validated performance metrics, their suitability maps showed major qualitative differences that required validation based on local expert knowledge. Additionally, our findings revealed varying importance of LULC and bioclimatic variables for different snail species at different spatial scales. Finally, we found that models using publicly-available data predicted snail distribution with comparable AUC values to models using expert-collected data. This work serves as an instructional guide to SDM methods that can be applied to a range of vector-borne and zoonotic diseases. In addition, it advances our understanding of the relevant environment and bioclimatic determinants of schistosomiasis risk in Brazil.

## Introduction

Species distribution models (SDMs) have become increasingly popular tools in the field of disease ecology to profile transmission risk for vector-borne and zoonotic diseases, i.e., diseases whose transmission involves a non-human host or vector species, such mosquitoes (malaria, dengue, Zika), flies (leishmaniasis, sleeping sickness), ticks (Lyme disease), triatomine bugs (Chagas disease), and snails (schistosomiasis, fascioliasis). By using presence data of non-human hosts and remotely-sensed data of potential environmental covariates, SDMs are correlative models that can predict species habitat suitability across areas not sampled by field collection programs (1–3). These models are typically used to create high-resolution maps of inferred species distribution across a geographic area of interest, which can reflect areas where disease transmission may be possible. In combination with other processes that influence transmission, such as reservoir host distributions or other disease exposure variables, these predictions can directly inform the understanding of the pathogenic landscape of environmentally-mediated diseases (4).

SDMs are a powerful tool applied in a number of fields, including disease ecology (5,6), epidemiology (7), and conservation (8,9), among many others. Species distribution modeling works by using presence/absence species data to identify covariates that are predictive of a species presence. Because true absence data are not typically available, SDMs often use “background” or “pseudo-absence” data to simulate locations where an organism could have been sampled but was not (10,11). SDMs use various machine learning methods to identify a suite of covariates that can accurately predict the presence or absence of the organism in geographic space, using flexible functional relationships between predictors and responses that can include nonlinearities and interactions (2,3). Model inputs can vary in spatial and temporal resolution and extent. Many algorithms are available for model training and testing, and they differ in how they handle covariate-outcome relationships (12). SDMs are cross-validated by leaving out part of the data in model training in order to assess model performance on out-of-sample data, often performed in a spatially-structured way (13). The outputs of interest include geographic maps of species presence suitability, lists of variables selected as important predictors, and the functional forms of relationships between predictors and presence. A glossary of terms and concepts central to the SDM literature are summarized for reference in **Table 1**.

**Table 1.** Glossary of terms and concepts central to SDM methodology.

| **Term/concept** | **Summary** | **Citation** |
| --- | --- | --- |
| Presence records | Observed presence of a species, usually with latitude, longitude, and date. | (33) |
| True absence records | Observed absence of a species, also with latitude, longitude, and date. Often difficult to obtain with certainty. | (34) |
| Pseudo-absence / background records | Locations drawn from the landscape of interest where an organism could have been sampled but was not. Could by chance include presence areas. | (10) |
| Thinning | Reducing a set of records (presence, absence, background) so only one record is retained for each grid cell. | (35) |
| Environmental covariates | Variables hypothesized to impact species presence, such as temperature, precipitation, or land-use type. Often remotely-sensed raster images. | (33) |
| Multicollinearity | When predictor variables are correlated to one another. Can potentially result in misidentification of relevant predictors, their importance, or their relationship with the outcome. | (36) |
| Resolution size | Size of the grid cells at which probabilities are predicted. All predictor variables need to be input into the model with the same resolution. | (27) |
| Geographic extent | The geographic area of interest for which probabilities are estimated. | (33) |
| Model type | Choice of statistical or machine learning algorithm. | (12) |
| Cross-validation / spatial cross-validation | Validation technique that repeatedly leaves out parts (i.e., “folds”) of the data in model training in order to assess performance on out-of-sample data. Spatial cross-validation separates the folds by spatial clusters. | (13) |
| Discrimination | The ability for models to distinguish between presence records and absence/background records. Often estimated by AUC values. | (37) |
| Sensitivity | The proportion of presences correctly identified as presences. Models with high sensitivity tend to build prediction maps that look more full. | (37) |
| Specificity | The proportion of absences/backgrounds correctly identified as absences/backgrounds. Models with high specificity tend to build prediction maps that look more sparse. | (37) |
| ROC-AUC | A measure of discrimination that weights the false positive rate (i.e., 1 - specificity) versus sensitivity across all possible thresholds. A value of 1 indicates perfect discrimination and 0.5 or less indicates performance is no better than random. | (37) |
| TSS | Sensitivity + specificity - 1 (i.e., values of zero or less indicate the performance is no better than random). A measure of discrimination designed to be less sensitive to species prevalence values than ROC-AUC. | (30) |
| Calibration | The degree to which the observed proportion of presences in a grid cell equates to the model estimated probability. Often evaluated with a calibration graph. | (38) |
| Variable importance | Measurement to estimate how much each covariate contributes to model performance. SHapley Additive exPlanations (SHAP) are a particularly useful method because they are model agnostic. | (39) |
| Partial dependence plots | Line plots that depict the marginal effect each predictor has on suitability probabilities. | (40) |

Increased access to large-scale, remotely-sensed environmental data (14,15) and species presence databases (16), such as the Global Biodiversity Information Facility (17), has spurred rapid expansion of these methods. Further, recent decades have brought rapid development of statistical models and machine learning algorithms that can be applied to species distribution models, such as regularized regression (18), decision tree (19), Bayesian (20), neural network (21), and ensemble methods (22), among many others. Although many machine learning methods have grown in popularity due to their flexibility, ability to model covariate interactions, and increasing accessibility in common programming languages like R, no single method has fully eclipsed its counterparts (12,23). There has been additional SDM methodological development, including optimization of sampling techniques for “background” or “pseudo-absence” points (11,24), increased rigor for input variable selection (25,26), investigation on resolution size (27), defense of spatial cross-validation techniques (13), integration of ecological theory (28,29), development of gold-standard model evaluation measures (30), and updated guidelines for method-specific reproducibility standards (31,32).

However, many of these new methods have not been recently documented, especially not in a cohesive, accessible manner for scientists new to SDMs or those interested in adopting new methods (41). To our knowledge, there has not been an analysis comparing machine learning algorithms, data sources, and geographic extents in combination and assessing the consequences for presence probabilities and covariate relationships. We hypothesize that algorithm performance will vary across geographic scales given differences in model structure, such as ability to handle covariate interactions and potential to overfit (12). Additionally, there are very few analyses that directly compare the effects of using GBIF presence records versus records from expert-executed field collection programs. Given the known spatial bias in GBIF data, we ask how well GBIF data can approximate predictions created from expert-collected data sources (42,43). Finally, although there has been discussion on the effect of resolution size (27), there has been limited discussion on how SDM performance varies across areas of differing geographic extent when resolution size is held constant.

In an effort to answer these methodological questions in a biologically and epidemiologically relevant study system, we will use the intermediate hosts of *Schistosoma mansoni* Sambon, 1907—*Biomphalaria* (Preston, 1910) snails—as a case study. Simultaneously, we will make substantial contributions to knowledge on predicting schistosomiasis risk in Brazil. Schistosomiasis is a debilitating parasitic disease caused, in Brazil, by *S. mansoni*, a parasite that relies on both freshwater *Biomphalaria* snails and human beings to complete its life cycle (44). In Brazil, approximately 6 million people are infected and 25 million live in areas where they are at risk of infection (45). The disease predominantly impacts poor communities dependent on open water sources for occupational activities or other components of daily life (46,47). More recently, schistosomiasis transmission has also been recorded in urban and peri-urban areas, impacting people who are either without access to basic sanitation services or whose sewage systems overflow in times of heavy rainfall (48,49).

Because *Biomphalaria* freshwater snails are obligate intermediate hosts of *S. mansoni* parasites, SDMs of the non-human hosts of schistosomiasis allow us to predict areas of suitable snail habitat where transmission may be possible. There are three competent *Biomphalaria* snail hosts in Brazil: *Biomphalaria glabrata* (Say, 1818), *Biomphalaria tenagophila* (D’Orbigny, 1835) and *Biomphalaria straminea* (Dunker, 1848). We restrict our main analyses to *B. glabrata* and *B. tenagophila* for ease of interpretation and include *B. straminea* findings in the supplement. Of the three, *B. glabrata* is the species most susceptible to the parasite and therefore of high public health importance, and *B. tenagophila* was the only species that had sufficient GBIF data to compare to expert-collected data. Because snails are ectotherms (i.e., their body temperature is dependent on their environment), their reproduction, survival, and dispersal are strongly affected by their surrounding temperature (50). The snails live in slow-moving freshwater, including permanent and temporary sources, which are both influenced strongly by precipitation and drainage patterns (51). Land-use and land-cover (LULC) characteristics affect snail presence through multiple pathways, including affecting temperatures through changes in tree canopy and vegetation cover and influencing water patterns through deforestation and agriculture (52). Finally, chemical factors and soil properties—such as pH and soil water content—are known to impact the survival of *Biomphalaria* snails, due to their impact on freshwater quality (53).

SDMs capture the snails’ biological relationships to these environmental factors and build predictive risk maps that can help to target disease intervention efforts such as mass drug administration (54). There have been multiple studies using SDMs to predict suitable snail habitat across multiple geographical scales in Brazil, from national (55–57) to sub-national analyses, including those specific to areas within Pernambuco (58), São Paulo (59), and Minas Gerais (60,61). However, all of these analyses test only MaxEnt models, with the exception of Guimarães et al., 2009 who used an indicator kriging procedure (60). Moreover, with the exception of Palasio et al., 2021, the quality and quantity of accessible, remotely-sensed environmental data has grown substantially since time of publication (59). Finally, our group has collected a large dataset of presence records throughout Brazil that reflect best expert knowledge of the constraints on snail habitat, presenting an alternative to publicly available GBIF presence data. Therefore, *Biomphalaria* snails in Brazil provide a ripe opportunity to compare and contrast current methods on SDMs, providing a rare comparative case study to guide SDM approaches for disease ecology and contributing updated risk models that can guide Brazil’s schistosomiasis elimination efforts (62).

We compare multiple combinations of SDM methods—three machine learning algorithms, two data sources, and three geographic extents—and assess the consequences for suitability probabilities and covariate relationships of two snail species. We address the questions: How do statistical/machine learning models compare depending on research question or application of interest? How do model accuracy, variable importance, and geographic predictions vary across spatial scales? How does model performance compare using expert-collected data versus publicly-available data?

## Methods

### Species data and background sampling

We acquired *B. glabrata* and *B. tenagophila* presence data from two main sources (1) an ongoing, Brazil-wide field program supported by multiple government-funded groups across Brazil, including the Coleção de Malacologia Médica, Fundação Oswaldo Cruz (CMM-Fiocruz) and the Coordination for Disease Control of the State Health Secretariat of São Paulo (CCD-SP) (63–70) and (2) the Global Biodiversity Information Facility (GBIF), a database of publicly available presence records commonly used to build SDMs (16).

The Brazil-wide field collection program, hereafter referred to as the expert-collected dataset, consisted of 11,299 total snail records that spanned 1992–2019 and included 25 species. As part of national efforts to control schistosomiasis, the Brazilian Ministry of Health has approved routine collection and monitoring of *Biomphalaria* snail species. Geographical coordinates of each collection site were acquired with a Garmin eTrex GPS device and species identification was done using morphological and molecular tools. Prior to model input, all records were spatially filtered such that only one presence record was retained for each 1km grid cell (i.e. “thinned to 1km”) to minimize pseudo-replication and oversampling bias (35). After each species was separately thinned to 1km, the dataset was reduced to 576 records of our snail hosts of interest: 305 *B. glabrata* and 271 *B. tenagophila* presence points (**Fig 1**, **Table 2**). *B. straminea* data quantity and distribution information can be found in **Supplementary Table 1** and **Supplementary Fig S2**.

**Fig 1:**
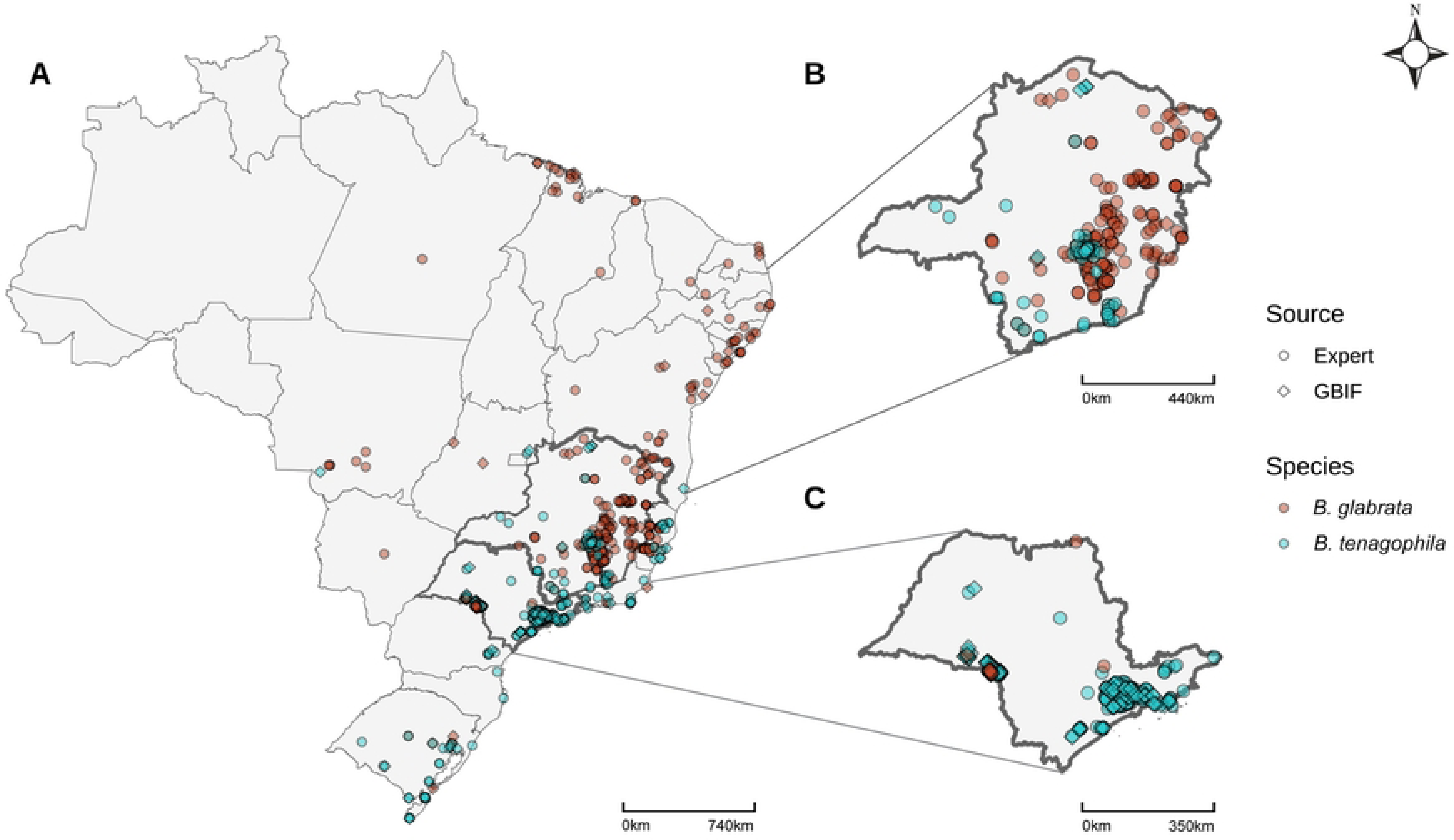
*Biomphalaria* presence points by species (color) and source (shape), thinned to 1 km. A) National, B) Minas Gerais, C) São Paulo

**Table 2.** *Biomphalaria* presence point quantity by species, scale, and source, thinned to 1 km.

| Species | Scale | Source | Presence point quantity | Proportion of national data |
| --- | --- | --- | --- | --- |
| *B. glabrata* | National | Expert+GBIF | 327 | 1 |
|  | Sudeste | Expert+GBIF | 244 | 0.74 |
|  | Minas Gerais | Expert+GBIF | 200 | 0.61 |
| *B. tenagophila* | National | Expert+GBIF | 358 | 1 |
|  | Sudeste | Expert+GBIF | 318 | 0.89 |
|  | São Paulo | Expert+GBIF | 234 | 0.65 |
|  | São Paulo | Expert* | 134 | NA |
|  | São Paulo | GBIF* | 115 | NA |

\*Source comparisons only made for *B. tenagophila* in São Paulo (the only scale where data quantities were comparable and sufficient).

To compare model performance between expert-collected and publicly available GBIF data and to create a background dataset (more below), we constructed a GBIF dataset by searching Brazil for all species included in the expert-collected dataset and records of all freshwater animals found in South America, as defined by the International Union for Conservation of Nature (71). This resulted in a total of 74,960 records that spanned 1985–2020, included over 2,000 species, and reduced to 165 records of our snail hosts of interest—29 *B. glabrata* and 136 *B. tenagophila*—post thinning. Our inclusion criteria for GBIF records were (i) year was between 1985–2020, (ii) latitude and longitude each included at least three decimal places and (iii) basis of record excluded “fossil specimen” and “machine observation” to ensure that the record was field-collected at the latitude and longitude reported and was identified by a human. For our snail hosts of interest we also required a complete species taxonomic identification. We limited our comparison of expert-collected versus GBIF data to *B. tenagophila* in São Paulo due to lack of sufficient data availability in other areas.

Given a lack of true absence data, we constructed a background dataset of freshwater animals across Brazil as our comparison group, thereby representing the freshwater landscape in which snails could plausibly be sampled. Species distribution models are often constructed using presence data only, without data on true absences of the species. To do so, models calculate the probability of species presence relative to the presence of a set of comparable species, assuming that the two have similar probabilities of being sampled given that they occur. In this way, “pseudo-absence” or “background” points aim to control for sampling effort to capture the relationships with environmental covariates that distinguish the presence of the species of interest from that of others (10). The extent of the background dataset should be chosen to represent the environmental variation of the study area (10). Our background dataset was a combination of (1) the remaining expert-collected data after excluding our three species of interest (4.8%) and (2) the publicly-available GBIF data described above (95.2%), which included a total of 2,091 freshwater animal species and 77,785 presence records. Each background dataset was built by sampling two times the number of presence data points for each model (i.e., a model with 100 presence points was given 200 background points): this ratio was selected to balance the sample between groups (72), while providing sufficient data to represent all environments and promote model convergence (11,73). Background points were sampled without replacement across a probability distribution that maintained the frequency of background points per 1km grid cell. Therefore, we retained a maximum of one record per grid cell, generating a “background mask” that helped address sampling bias concerns (10,74).

### Environmental data and multicollinearity analysis

We used high-resolution (1km) climate, hydrology, soil property, and land-use/land-cover (LULC) data to describe the environmental conditions associated with each species presence record and background sample. We limited the number of covariates to variables previously found to impact snail presence for ease of interpretation and comparison between model design choices (36). Climate data were obtained from CHELSA (version 2.1), a high resolution (1km^2^) global downscaled climate data set (75). Four climatology variables, averaged over thirty years (1981–2010), were included in the analysis: temperature seasonality (bio4), mean temperature of coldest quarter (bio11), mean precipitation of wettest quarter (bio16), and mean precipitation of driest quarter (bio17). Hydrology data (height above nearest drainage—HND—and soil water percentage) were obtained from the Merit Hydro data (76) and OpenLandMap Soil Water Content (77), respectively, and soil property data (pH and clay) was obtained from OpenLandMap Soil pH in H_2_O (78) and OpenLandMap Clay Content (79), respectively. Because hydrology and soil variables were measured at finer spatial resolution than the climate data, we scaled them up to the maximum value (HND) or mean value (water content, pH, clay content) for each 1km^2^ grid cell. Finally, our two LULC covariates—distance to high population density and proportion of temporary crop cover during the year of sampling—were constructed from WorldPop (80) and MapBiomas (81), respectively. High population density was defined as a 1km grid cell with a density of at least 1500 inhabitants per km^2^, per the World Bank definition (82). Proportion of temporary crop cover was defined—in natural areas—as farming areas where it was not possible to distinguish between pasture and agriculture and—in urban areas—as areas of urban vegetation, including cultivated vegetation, natural forest, and non-forest vegetation (81). We selected these two LULC variables based on our team’s on-the-ground knowledge of snail presence (83). In total, we provided our models with 12 environmental covariates (**Supplementary Fig S1**), none of which had pairwise Pearson correlation coefficients above 0.7 with any of the other covariates (36). Although our studied models can handle multicollinearity when calculating probabilities, collinear variables obscure the variable importance and partial dependence plot interpretation (36).

### Geographic extent

To investigate model performance across varying geographic extent, we created models spanning national, regional (Sudeste, composed of four states: Espírito Santo, Minas Gerais, Rio de Janeiro and São Paulo), and state (Minas Gerais and São Paulo) extents in Brazil. The region and states of interest were chosen based on the quantity of data available to input into the models. Past studies have shown that model performance substantially declines with fewer than 30-50 presence records (84,85). We selected only states with greater than 100 presence records for a species of interest: Minas Gerais for *B. glabrata* and São Paulo for *B. tenagophila* (**Table 2**).

### Statistical model type

To compare between machine learning modeling methods, we built three model types: Maximum Entropy (MaxEnt), Random Forest (RF), and Boosted Regression Tree (BRT). We chose these three model types due to both their popularity in the literature and their consistently high performance (12). MaxEnt has long been a well-established method for presence-only applications (86), while model types such as RF and BRT have gained more recent popularity (41). All models were built using the R program (version 4.2.2).

MaxEnt uses a maximum-entropy approach to estimate a species’ relative probability distribution in response to environmental covariates (18). MaxEnt models create smooth fitted curves, which can facilitate straightforward ecological interpretation (10). The degree to which this “smoothness” is enforced can be controlled through choice of regularization settings and by which feature types are provided, where options include linear, quadratic, hinge, threshold, and product features (10). Product features are equivalent to interaction terms in regression, and they allow for limited interactions between covariates (10). We allow MaxEnt all five of these options and use the *trainMaxNet* function from the *enmSdmX* package, which includes an L1 regularization feature (87).

On the other hand, RF, BRT, and other tree-based methods provide enhanced flexibility that allow for automatic fitting of precise interactions between the environmental covariates (88). RF models take bootstrap samples from the training data and fit a decision tree to each sample (73). These individual trees can have high variance (i.e., depend heavily on the training data), but have strong generalizability when averaged together to make a prediction over all fitted trees (88). RF models use random subsets of the available predictor variables (parameter *mtry*) on each decision tree split, which results in decorrelated trees and subsequently improves model performance (73,89). Due to its relative ease of implementation and conceptual simplicity, RF has become a common SDM approach (12). However, RF models have the potential to overfit, especially when provided data with high class imbalance (73). We use the *trainRF* function from the *enmSdmX* package (87), which is a wrapper of the *randomForest* function from the *randomForest* package (90).

BRT is similar in structure to RF, but the decision trees are recursively updated as the algorithm learns. During each step of the learning process, BRT fits new trees to the residuals for the previously fitted trees, which allows the algorithm to improve on the observations that are not yet predicted correctly (19). We use the *trainBRT* function from the *enmSdmX* package (87), which is a wrapper of the *gbm.step* function from the *dismo* package (91). Similar to RF, BRT also has the potential to overfit to training data but can better handle class imbalance and missing data due to its additional hyperparameters (19). While these hyperparameters make BRT the most flexible model of the three included in our analysis, they require an additional tuning step that can be computationally expensive (19). As of now, no one model type has fully eclipsed the others as the SDM standard, but tree-based methods have been shown to improve performance in multiple settings (12).

### Model evaluation

Our goals in model evaluation were first to assess the accuracy of each model in classifying presence versus background (how well does each model classify snail distribution?), second to compare model accuracy among methods (which machine learning approach represents the data best?), third to assess the importance of different environmental covariates and the shapes of their relationships with presence (what environmental characteristics are positively or negatively associated with the observed snail distribution?), and fourth to compare this variable importance and functional form among model methods (are the relationships between predictors and snail presence consistent among models?). Before quantifying accuracy, we first assessed model biological realism qualitatively by using expert opinion to visually compare maps produced by giving models the full set of records available for each species at the scale of interest. Our group of experts consisted of scientists from CMM-Fiocruz and CCD-SP who have studied and organized field collection of *Biomphalaria* snails in Brazil for over three decades. Second, we assessed accuracy using four out-of-sample model performance metrics, as described below, calculated through ten-fold spatial cross-validation (a process where folds are divided in space instead of through random sampling, which can inflate SDM performance measures due to spatial autocorrelation of environmental covariates (13)). We determined the ten spatial folds using a k-means clustering algorithm where the size of folds was allowed to vary to prioritize the degree of spatial separation. Each fold was required to have at least one presence and one background point.

To determine each model’s discrimination ability, we calculated sensitivity, specificity, the area under the receiver operator characteristic curve (ROC-AUC), and true skill statistic (TSS). Sensitivity is the proportion of presences correctly identified as presences, and specificity is the number of background points correctly identified as background records. ROC-AUC measures the false positive rate (i.e., 1 - specificity) versus sensitivity across all possible thresholds (37). An AUC value of 1 indicates perfect discrimination and 0.5 or less indicates the performance is no better than random). We allowed AUC threshold values to vary across each fold for each model (92). TSS is defined as *sensitivity + specificity - 1* (i.e., values of zero or less indicate the performance is no better than random) and is designed to be less sensitive to species prevalence values (30). Given our interest in comparing each of our models’ ability to distinguish relative suitability of sites, output suitability probabilities were scaled such that all distributions ranged from 0 to 1 (38,85). Calibration is the degree to which the observed proportion of presence records in a grid cell equates to the model estimated probability (i.e., 60% of grid cells predicted with a probability of 0.6 contain a presence record (38)). The main calibration evaluation technique is a calibration graph, which plots model probability estimates against the observed proportion of presences, and is predominantly used in studies with true absence data (38). Although not applicable for this analysis, there are other situations where the calibration of the model is an additional aspect that should be tested, such as when evaluating estimates of true prevalence (38).

Finally, partial dependence plots and variable importance measures were calculated across the ten folds for each model to investigate each covariate’s contribution to model accuracy and functional relationship with presence probability. Partial dependence plots (PDP) were drawn using the *pdp* R package and show the marginal effect of each predictor on model probabilities (93). Partial dependence plots allow for comparison of inferred relationships between covariates and occurrence probability with *a priori* knowledge of factors that drive snail ecological niche suitability. Variable importance measures were calculated using the *vi_shap* function from the *vip* package (94), which calculates SHapley Additive exPlanations (SHAP) variable importance values (a method of calculating how much covariates contribute to model predictions) (39,95). Notably, SHAP values are model agnostic and can estimate comparable values of variable contribution for both regression-based and tree-based methods (39).

## Results

Model types produce remarkably different national prediction maps for all species despite using the same presence and background records and environmental data (**Fig 2**; **Supplementary Fig S3**). Although probability prediction varies widely (**Fig 2**), spatially cross-validated AUC (**Fig 3**) and TSS (**Supplementary Fig S4**) values of national models do not substantially differ across model types. RF models tended to have somewhat higher sensitivity—they were more likely to accurately predict presence points—than MaxEnt and BRT across species (**Table 3**; **Supplementary Fig S4**). BRT models had higher specificity—they were more likely to accurately predict background points—than MaxEnt and RF for *B. glabrata* (**Table 3**) and *B. straminea* models (**Supplementary Fig S4**). When comparing de-identified national prediction maps, expert opinion selected BRT maps for both *B. glabrata* and *B. tenagophila* as best matching *a priori* knowledge of current suitable snail habitat.

**Fig 2:**
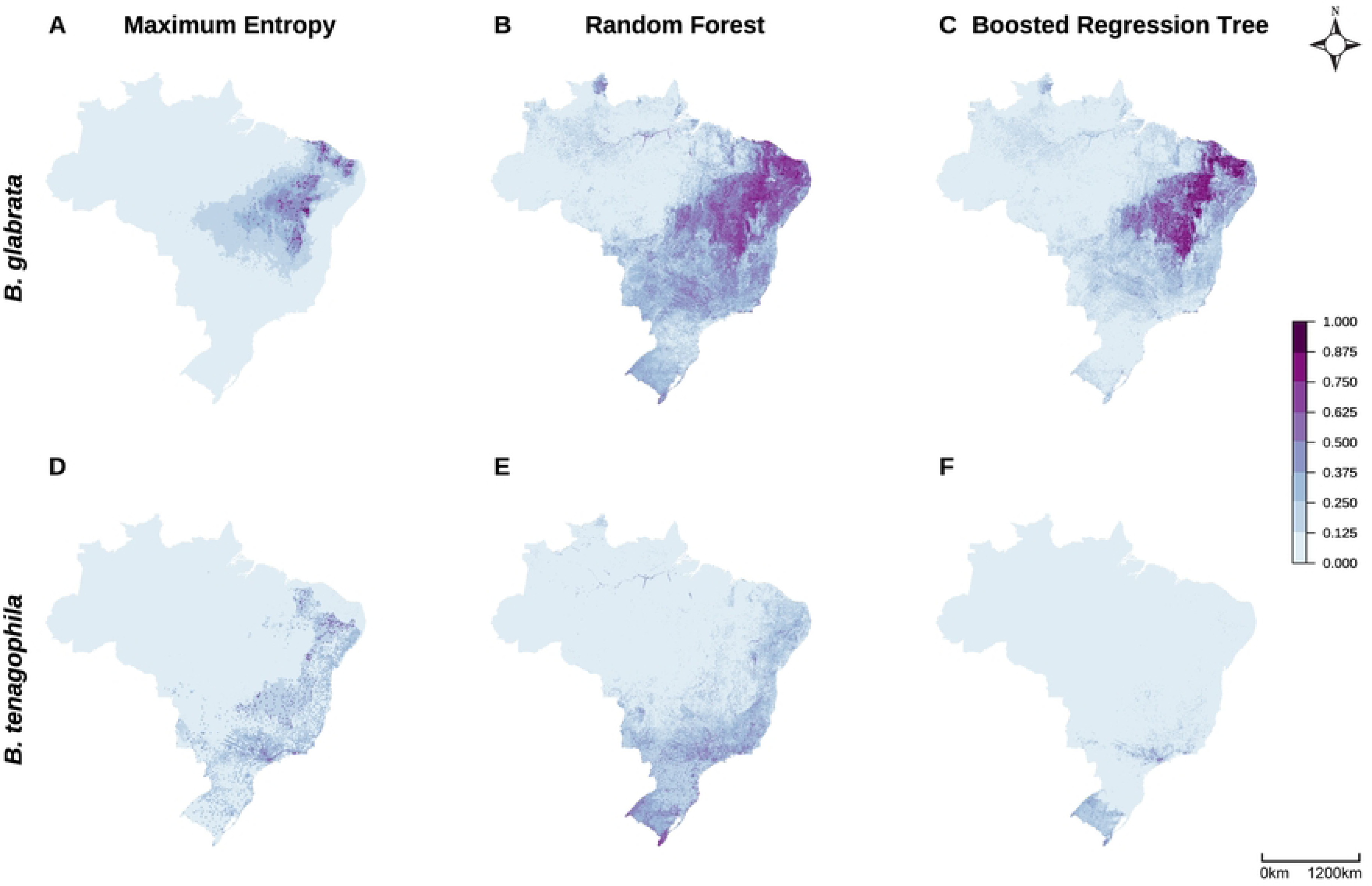
Large variation in snail suitability probabilities at a national scale. National prediction maps of *B. glabrata* (A–C) and *B. tenagophila* (D–F) suitability probabilities by model type (MaxEnt: A, D; Random Forest: B, E; Boosted Regression Tree: C, F) when models were provided the full set of species presence records available.

**Fig 3:**
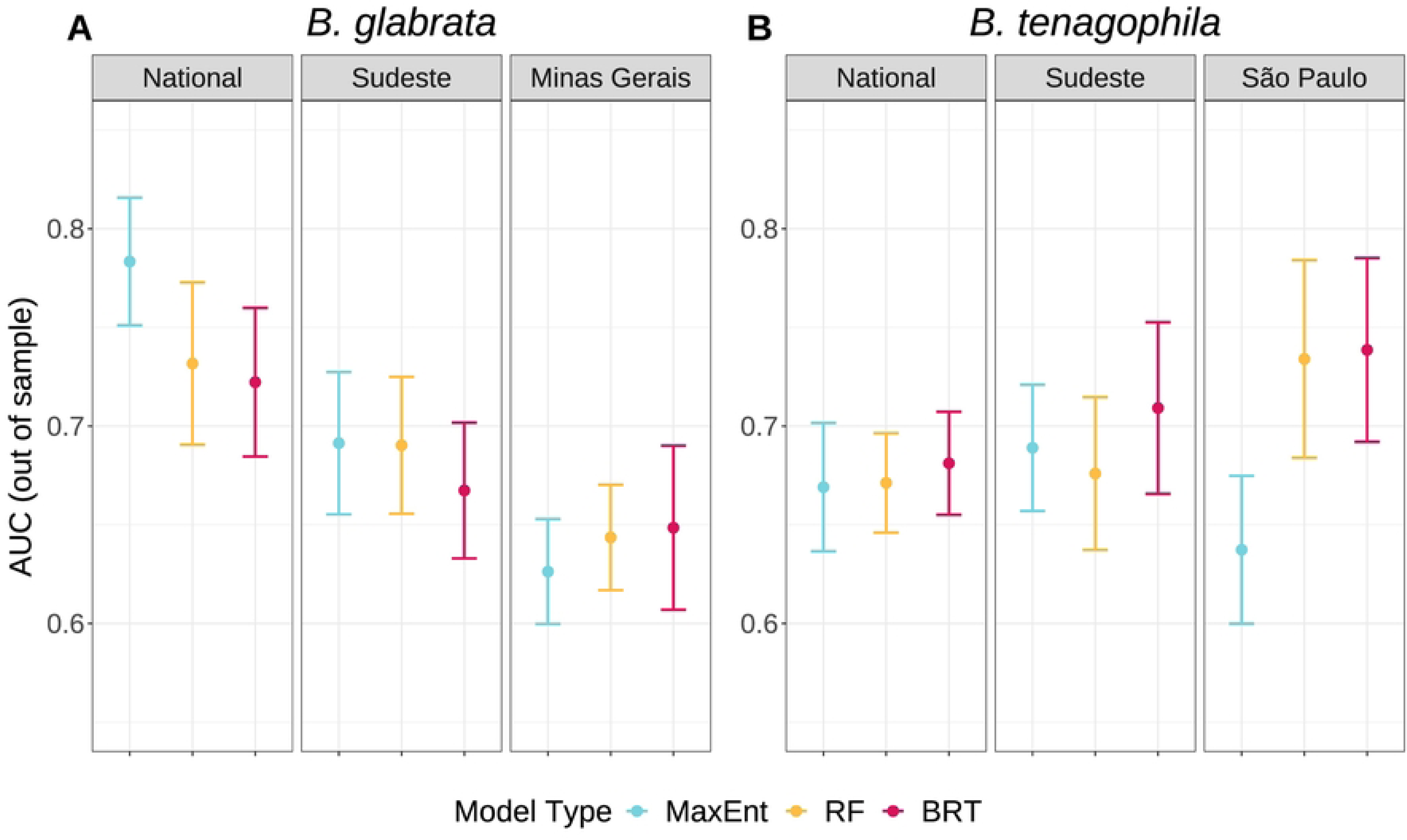
Scale and species drive SDM performance metrics more than model type. Plots of ten-fold spatially cross-validated, out-of-sample AUC values across species (A, B), scales (panels), and model types (colors). Plots display mean (point) and +/- standard error (error bars).

**Table 3.**
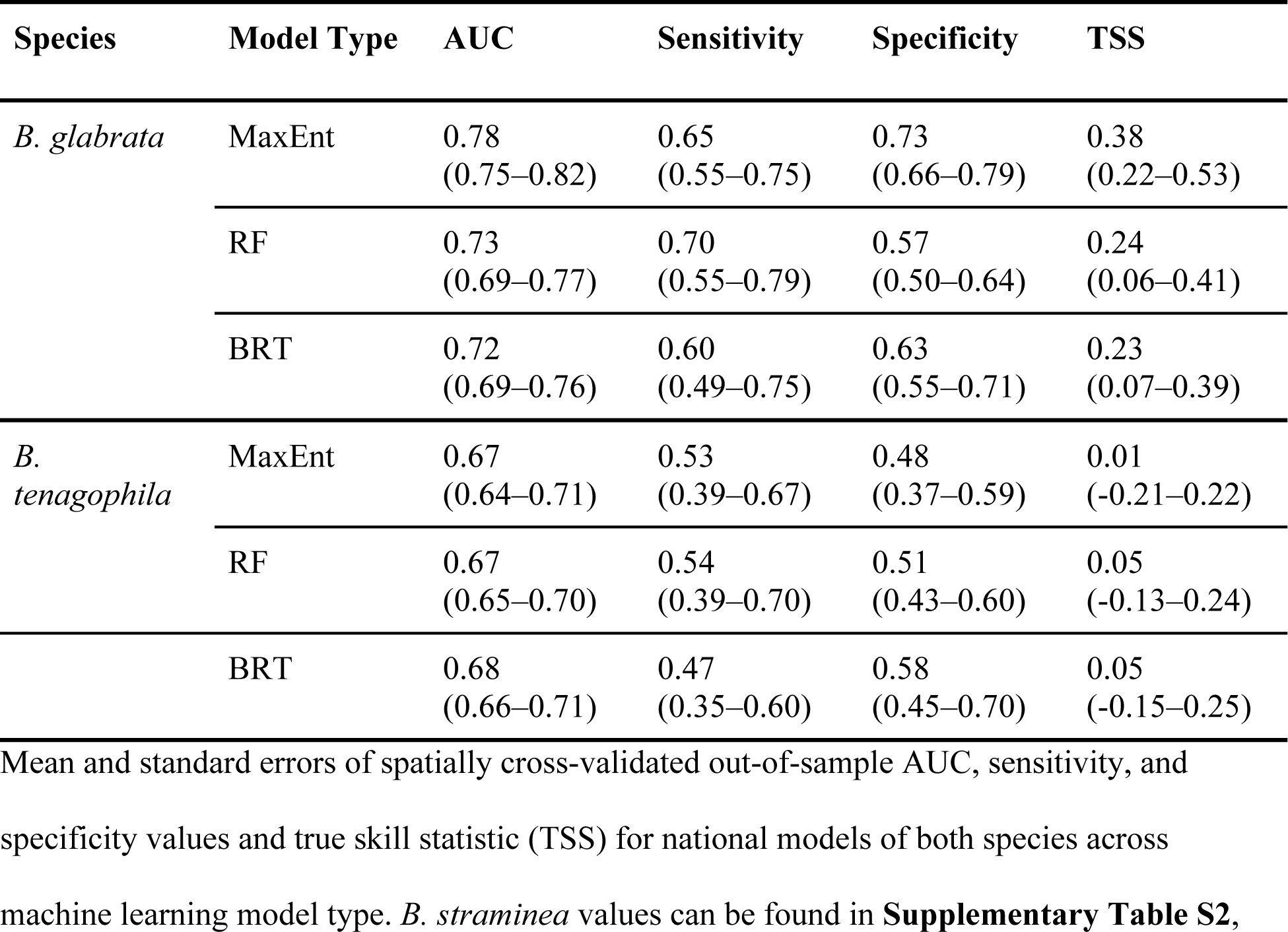
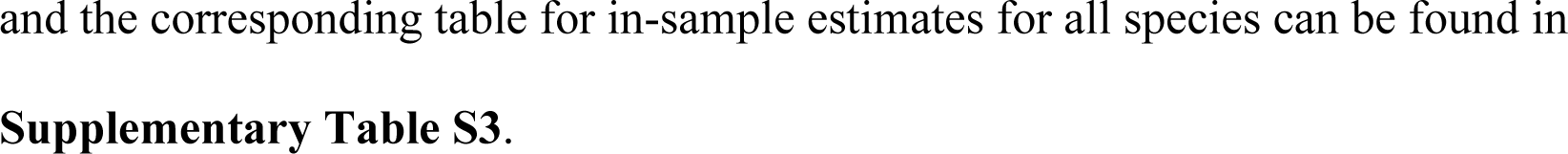
Model performance of national *Biomphalaria* snail models across machine learning model types.

| Species | Model Type | AUC | Sensitivity | Specificity | TSS |
| --- | --- | --- | --- | --- | --- |
| *B. glabrata* | MaxEnt | 0.78<br>(0.75–0.82) | 0.65<br>(0.55–0.75) | 0.73<br>(0.66–0.79) | 0.38<br>(0.22–0.53) |
|  | RF | 0.73<br>(0.69–0.77) | 0.70<br>(0.55–0.79) | 0.57<br>(0.50–0.64) | 0.24<br>(0.06–0.41) |
|  | BRT | 0.72<br>(0.69–0.76) | 0.60<br>(0.49–0.75) | 0.63<br>(0.55–0.71) | 0.23<br>(0.07–0.39) |
| *B. tenagophila* | MaxEnt | 0.67<br>(0.64–0.71) | 0.53<br>(0.39–0.67) | 0.48<br>(0.37–0.59) | 0.01<br>(-0.21–0.22) |
|  | RF | 0.67<br>(0.65–0.70) | 0.54<br>(0.39–0.70) | 0.51<br>(0.43–0.60) | 0.05<br>(-0.13–0.24) |
|  | BRT | 0.68<br>(0.66–0.71) | 0.47<br>(0.35–0.60) | 0.58<br>(0.45–0.70) | 0.05<br>(-0.15–0.25) |

and the corresponding table for in-sample estimates for all species can be found in **Supplementary Table S3**.

Compared to these national models, model accuracy remained consistent at smaller geographic scales for *B. glabrata* and increased at smaller geographic scales for *B. tenagophila* (**Fig 3**; **Supplementary Fig S4**), as measured by spatially cross-validated out-of-sample AUC, TSS, sensitivity, and specificity. However, when testing models fit to national-scale data at predicting state-level occurrences, all models for both species produced lower in-sample AUC values than state models (**Supplementary Table S4**). State models also generally produced higher in-sample sensitivity and specificity values but the nationally-fit models occasionally produced higher sensitivity values (i.e., sometimes the nationally-fit models were able to correctly identify presence points that the state-specific models missed). Differences in predictive accuracy between models trained on state versus national data when tested on state data occurs due to differences in predicted suitability maps, which are visually apparent (**Fig 4, Supplementary Figs S5 and S6**).

**Fig 4:**
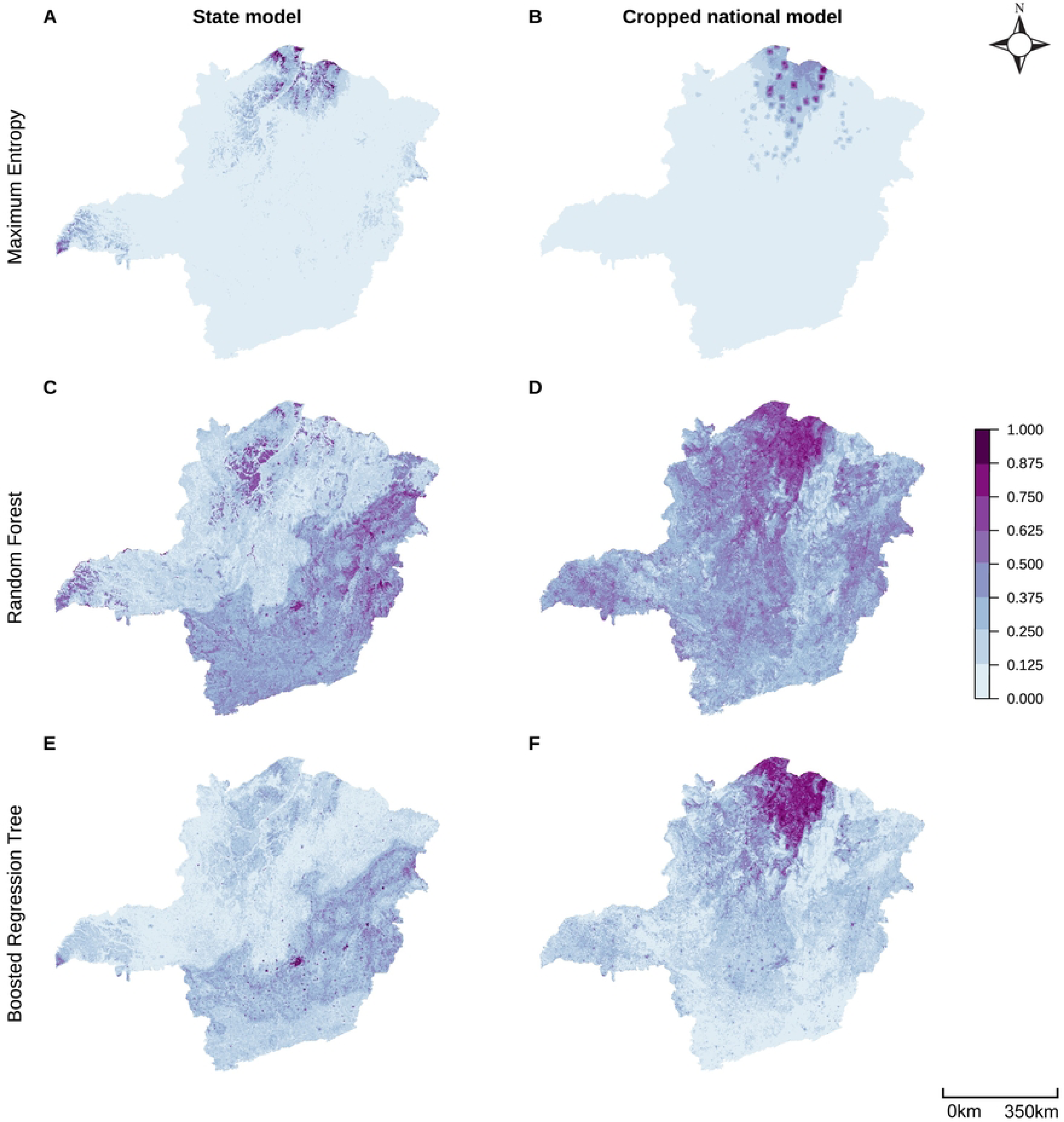
State and national models produce substantially different state-level prediction maps. Minas Gerais prediction maps of *B. glabrata* suitability probabilities by model type (rows) and model geographic extent (columns) when models were provided the full set of species presence records available at a given scale. Parallel prediction maps of *B. tenagophila* in São Paulo and *B. straminea* in Minas Gerais can be found in **Supplementary Figs S5 and S6**. Compared to national models (**Fig 2**), at smaller geographic scales it becomes more obvious that suitability probabilities can be highly localized, producing points of high suitability surrounded by areas with low suitability.

Despite similar overall accuracy across machine learning model types within geographic extents (i.e., MaxEnt national compared to RF national (**Fig 3**), the models infer strikingly different relationships between covariates and suitability probability, which imply distinct biological relationships. We use three specific examples to illustrate how responses differ across model types, spatial extents, and focal species, by comparing plots in each column of **Fig 5**. First, model types produce different curve shapes: MaxEnt often fits smoother or linear forms in comparison to the flexible, nonlinear shapes produced by RF and BRT, as illustrated by distance to high population density (**Fig 5A**). Second, functional forms vary across scales: both *B. glabrata* and *B. tenagophila* responses to soil clay percentage are directionally opposite at national versus state scales for all model types (**Fig 5B**). It is important to note that the range of environmental covariates may differ remarkably across geographic extents. Third, species differ in the functional forms: *B. glabrata* and *B. tenagophila* suitability both respond nonlinearly to temperature in the coldest quarter, but with different functional responses that vary between scales (**Fig 5C**). By contrast, other functional forms remain relatively consistent across species and scale, such as the response to distance to high population density (**Fig 5A**). These differences in inferred biological relationships highlight the potential pitfalls of using SDMs to extrapolate environmental suitability beyond the scope of the data, and of assuming generality from a single modeling approach.

**Fig 5:**
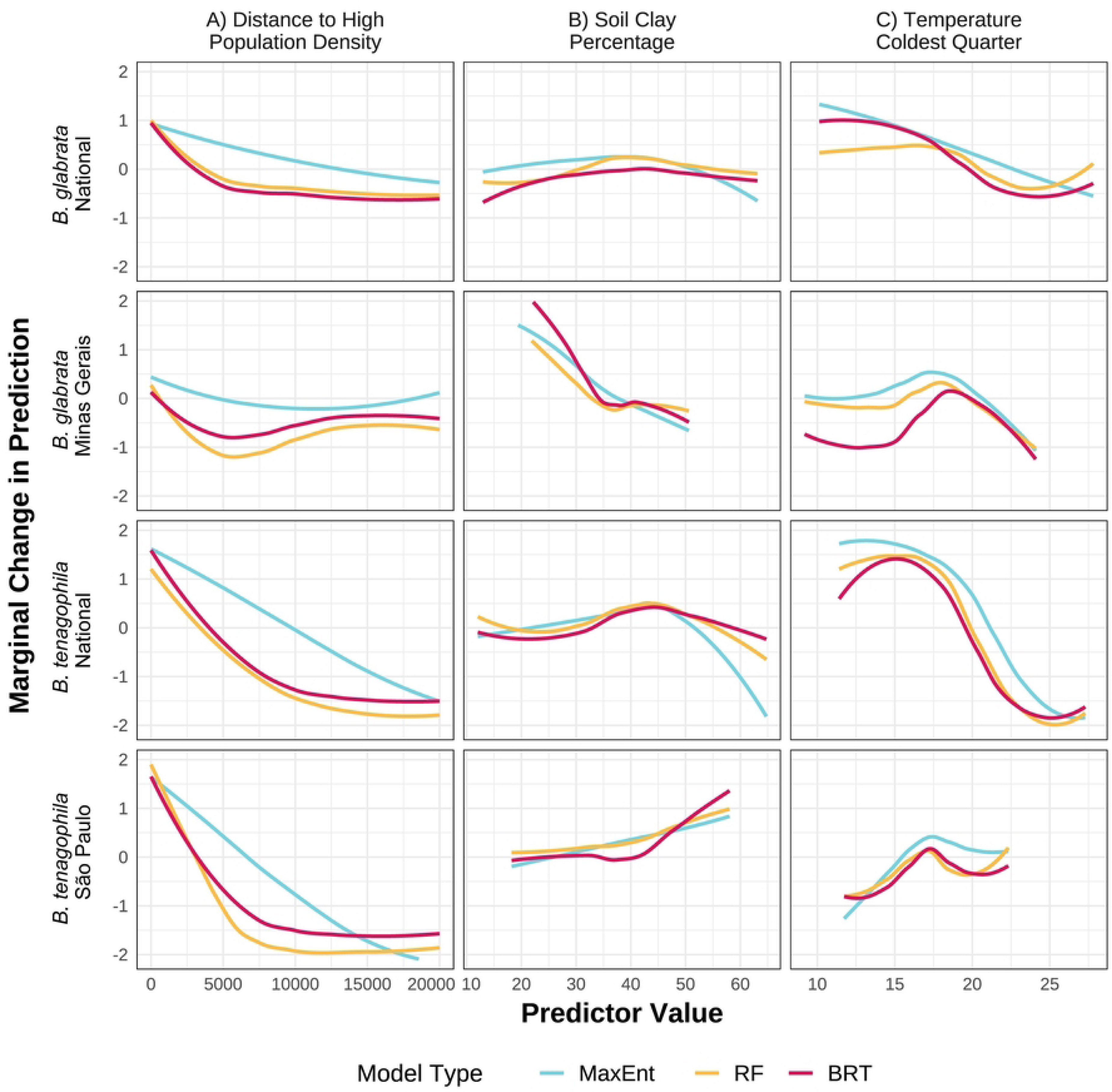
Examples of marginal effects of covariates on suitability probabilities that vary across model type (A), geographic scale (B), and species (C). Partial dependence plots for three covariates (columns) across model types (color), species (top two rows vs. bottom two rows), and scale (first row vs. second and third row vs. fourth).

Given the importance of understanding how *Biomphalaria* snails are responding to land use and land cover (LULC) change, we investigated how the relative importance of LULC variables changes with scale. We hypothesized that LULC variables would become increasingly important compared to climatic gradients at relatively smaller scales. Evidence for this prediction was mixed. Supporting this prediction, the relative importance of LULC variables increased consistently from national to regional to state scales for *B. tenagophila* models (**Fig 6B**). However, LULC variable importance for *B. glabrata* models (**Fig 6A**) remained more constant across scales and decreased at the state scale when using a MaxEnt model. Similar to *B. glabrata,* LULC variable importance for *B. straminea* models (**Supplementary Fig S7**) dipped in regional models and was equivalently high in national and state models.

**Fig 6:**
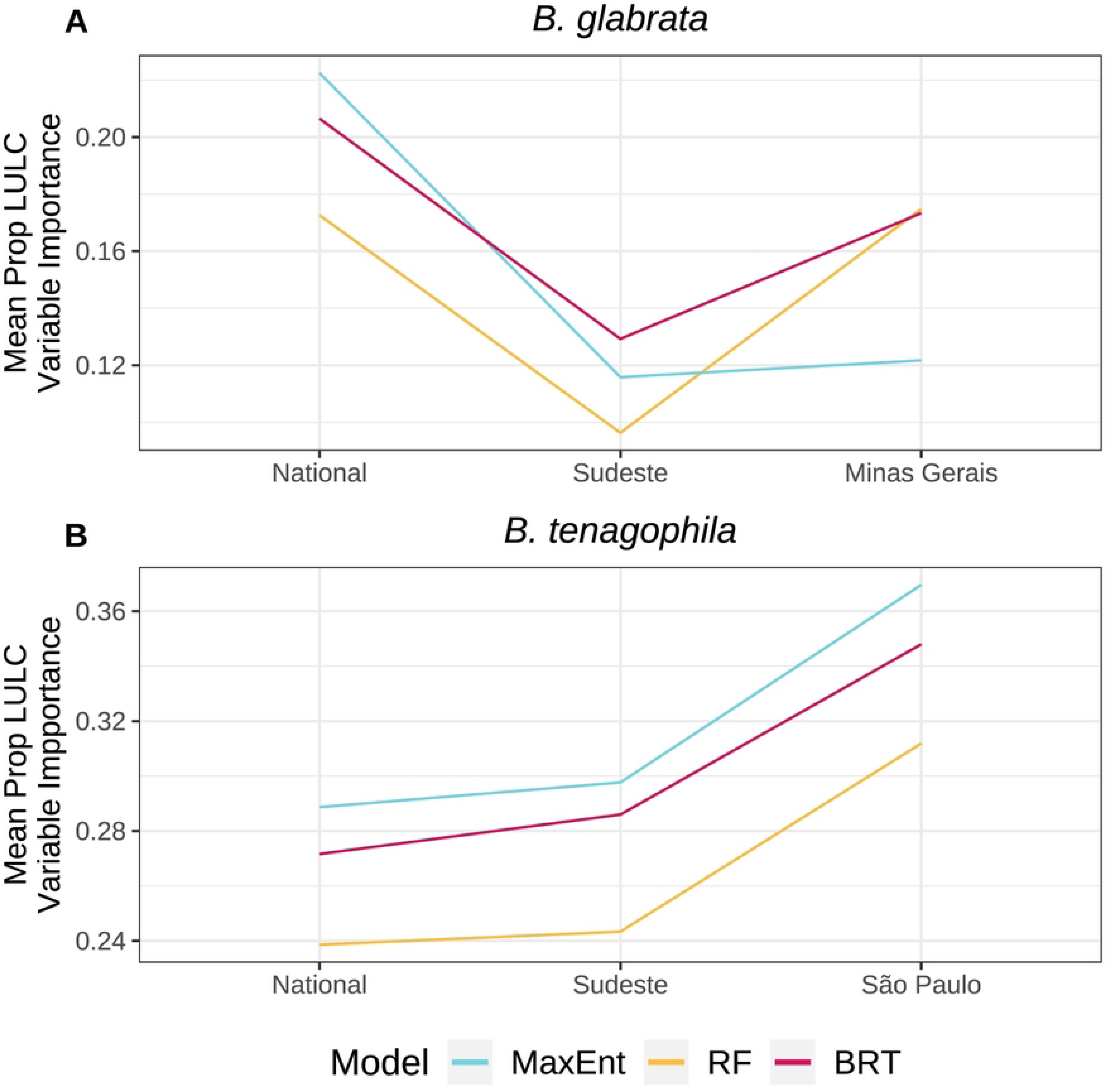
Variable importance of land use/land cover (LULC) variables can increase at smaller scales. Proportion of total variable importance averaged across all training folds attributable to distance to high population density and proportion of temporary crop cover. Displayed for all species (A vs. B) and model types (color).

To investigate impacts of using presence data from an expert-executed field collection program versus from a publicly-available species presence database, we constructed models using two distinct datasets: expert-collected and GBIF. As anticipated, each dataset produced distinct predictions of presence probability. Limiting these analyses to *B. tenagophila* in São Paulo, model accuracy was similar across both datasets (expert-collected mean AUC = 0.83, 10th/90th percentiles: [0.71, 0.95], publicly-available GBIF mean AUC = 0.79, [0.66, 0.93]), yet the prediction maps show substantial variation regardless of model type (**Fig 7**). Despite somewhat lower AUC values when the two datasets were combined (0.70, [0.57, 0.92]), experts judged the suitability maps as preferable when data from both sources is included, across all model types (**Fig 7C, 7F, and 7I**). Notably, the two data sets have different data quantities, with the expert-collected dataset (*n* = 169) containing more presence points than the GBIF dataset (*n* = 115). The same maps and AUC comparisons for models with data quantity held constant (*n* = 115) can be found in the Supplementary Materials (**Supplementary Fig S8**), with slightly more variation between model suitability maps but generally small changes to above results.

**Fig 7:**
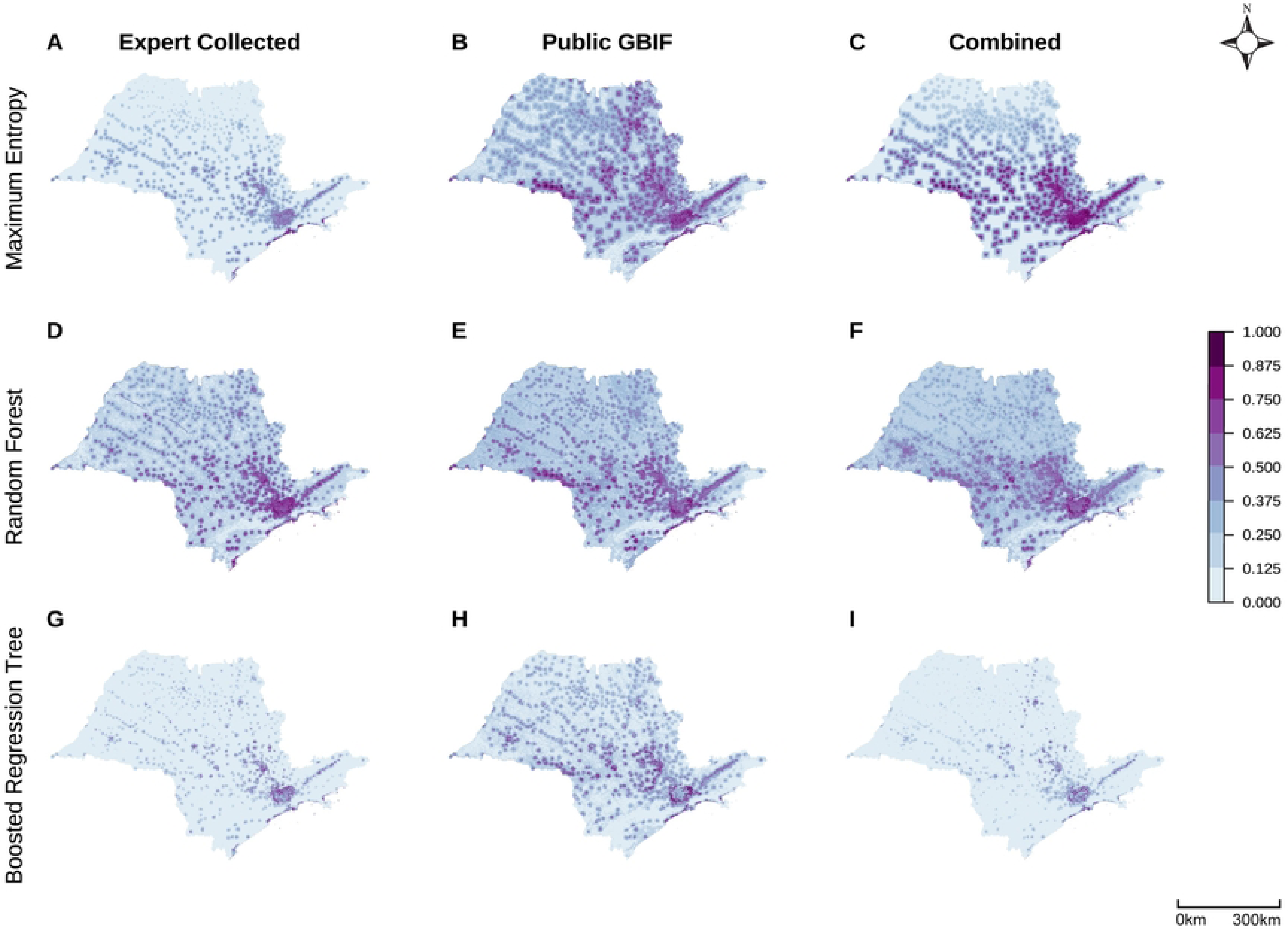
Expert collected and public GBIF data produce visually different suitability maps for *B. tenagophila* in São Paulo across model type. Predicted suitability maps with varying input data (columns) supplied to all model types (rows). Compared to national models (**Fig 2**), at smaller geographic scales it becomes more obvious that suitability probabilities can be highly localized, producing points of high suitability surrounded by areas with low suitability.

## Discussion

SDMs are increasingly used in disease ecology to understand environmental drivers of reservoir host or vector species distributions and to project how they might change with anthropogenic modification. We showed, by systematically comparing SDM approaches that employed different modeling techniques, spatial extents, data types, and species, that both the spatial predictions and the inferred relationships with environmental features can vary substantially across methods, even when performance measures (i.e., sensitivity, specificity, AUC, and TSS) are very similar.

A first important result is that even when given the same occurrence, background, and covariate data, the three model types produce remarkably different suitability maps despite similar accuracy. Although differences in spatially-cross validated mean AUC values were minimal when compared within geographic extents (i.e., MaxEnt national compared to RF national), we found that RF models tended to have higher sensitivity, producing more ‘dense’ maps of predicted suitable habitat, than MaxEnt or BRT across species and scales (**Table 3; Supplementary Fig S4**). On the other hand, BRT models tended to have higher specificity, producing more ‘sparse’ predictions, as compared to MaxEnt and RF across scales for *B. tenagophila* and *B. straminea* models (**Table 3; Supplementary Fig S4**).

Our analysis demonstrates the importance of individually investigating sensitivity and specificity (separate from AUC), especially if models are intended to inform disease control policy (96,97). If total elimination is of high priority, high sensitivity—the ability for models to accurately identify all presence locations—might be emphasized to safely capture all presence areas, with less concern for mistakenly implementing control interventions in places that actually contained only background records, which in this case would generally suggest using RF models for most species and scales (**Table 3**; **Supplementary Fig S4**). Alternatively, with more limited resources, policymakers might prioritize models with high specificity (i.e., the ability to accurately identify locations where the species is not expected), such as the BRT models at all scales for *B. tenagophila* and *B. straminea* (**Table 3**; **Supplementary Fig S4**). These models would minimize potential efficiency losses that could result from control programs deploying available resources in places that do not actually contain the species of interest. Finally, our experts consistently selected de-identified BRT models as producing maps that best aligned with their *a priori* knowledge of suitable snail habitat across multiple geographic contexts (national and São Paulo scales): these models tended to have higher specificity and lower sensitivity, making their occurrence predictions relatively more sparse. Overall, our findings align with previous comparisons of statistical model types in the SDM literature: MaxEnt, RF, and BRT can all produce high model performance measures, although which is the best can vary across species types (12,33). Therefore, we encourage modelers to use the suite of SDM resources (*dismo*, *enmSdmX*, etc.) to draft multiple models for their application and explicitly test which model type is best suited for their question, as detailed further below.

When prediction maps are used to inform intervention and/or funding decisions, significant differences in the suitability maps could warrant radically differing deployment of control strategies (54,96,97). Therefore, in addition to evaluating multiple model performance measures (AUC, sensitivity, specificity, TSS, etc.), it is crucial to leverage local ecological knowledge to assess the biological realism of each model’s predicted suitability map, as well as of the estimated ecological relationships derived from partial dependence plots (83). Other analyses have leveraged expert assessment of model outputs when AUC was unable to clearly rank models by performance (42). This aligns with the well-known but underemployed guideline that remotely-sensed, big data models need to be integrated with local, on-the-ground knowledge to create the best understanding of the system of interest (83).

Subtle differences in performance across scales suggest that the most relevant geographic extent may depend on the application and the relative distribution of data at different geographical scales; yet we also found that model performance could be high from national down to state scales. Comparing across geographic scales, spatially cross-validated AUC values decreased at smaller geographic scales for *B. glabrata* and *B. straminea*, but increased at smaller geographic scales for *B. tenagophila* (**Fig 3**; **Supplementary Fig S4**). This phenomenon can likely be attributed to the varying proportion of presence data for each species within each state (**Table 2**; **Supplementary Table S1**). While 89% of national *B. tenagophila* data is from within the Sudeste region and 65% is within São Paulo state, only 74% of national *B. glabrata* data is from the Sudeste region and 61% is from Minas Gerais. *B. straminea* had an even smaller proportion of total national data at the state level (**Supplementary Table S1**). Accordingly, we hypothesize that larger amounts of localized data for *B. tenagophila* Sudeste and São Paulo models improved model accuracy, while limiting the ability for national models to capture ecological heterogeneity across the entirety of Brazil. On the other hand, *B. glabrata* and *B. straminea* records are more widely distributed across the nation, allowing for improved national predictions, whereas the smaller data set from Minas Gerais limits the performance of state and regional models. When specifically aiming to create best predictions for small geographic regions, we demonstrate that locally-fit SDMs moderately increase model discrimination ability (**Supplementary Table S4**) and create maps with visually finer resolution predictions as compared to nationally-fit models (**Fig 4**; **Supplementary Figs S5 and S6**). However, when data are more uniformly distributed at the national scale, national scale models can be cropped to smaller scales relatively effectively, indicating that building national models can also be warranted when needed for large-scale applications or when investigating smaller geographic regions that have limited local data. A final key factor affecting choice of geographic extent is whether the aim is to identify covariate relationships specific to a geographic area of interest or to see generalized covariate relationships that span heterogeneous habitats and geographies, including ranges not yet observed in a given geographic region. This is particularly important when researchers aim to use SDMs to project species distributions under scenarios of future climate change, which include temperature and precipitation patterns not yet experienced in a given region.

Covariate relationships not only varied depending on geographic extent, but also by species and machine learning model used. Even for two snail species in the same genus, their responses to environmental covariates varied in both magnitude and direction (**Fig 5**), contributing to the large suitability map differences (**Fig 2**; **Fig 7**; **Supplementary Fig S3**). Compounding these true biological differences among species is the fact that different model structures produce differently shaped partial dependence plot curves, weighting interpretability versus flexibility and differentially favoring nonlinearity and interactions (10,86,88). For example, even when providing our MaxEnt models maximum flexibility in fitting the observed data, the resulting PDP curves still exhibit more limited shapes than RF or BRT. MaxEnt’s smooth curves offer simple, interpretable predictor relationships—potentially preferred for modelers whose primary interest is general mechanisms that underlie habitat suitability and/or ease of explanation for policymakers who need to make decisions with limited time (10). On the other hand, the hyper-flexible curves produced by RF and BRT (and other tree-based methods) can produce improved model performance and variable interactions, especially when models include suites of variables known to interact in nonlinear ways, such as temperature and precipitation or sets of LULC variables (19,73,88). If model classification ability is held at the highest priority and modelers believe it is ecologically feasible for predictors to have flexible relationships, partial dependence plots and the other model evaluation methods discussed here can assist in retaining clear model interpretation (39,95). Finally, we note that SDMs are correlative analyses. Therefore, modeled covariate relationships may not be directly related to species presence but with other environmental variables not included in the model. SDMs should be followed by causal analyses if the goal is to understand true causes of species presence.

LULC variables became proportionally more important for predicting *B. tenagophila* snail presence at smaller geographic scales as compared to bioclimatic variables. However, LULC variable importance remained relatively constant across scales for both *B. glabrata* and *B. straminea*. Given remotely-sensed bioclimatic variables predominantly change at larger spatial scales (i.e., they are highly spatially-autocorrelated), we expected that models of smaller geographic extent would rely more heavily on LULC variables, which contribute more localized variation (**Supplementary Fig S1**). This hypothesized effect may have been mitigated for two of the three snail species due to the fact that we held spatial resolution (1km^2^) constant over the three geographic extents. Other analyses varying resolution size have shown that biotic interactions dominate at local scales, while abiotic factors dominate regionally (27). Holding resolution constant likely allowed even national scale models to leverage localized variation derived from LULC variables.

Given an adequate number of presence points, publicly-available GBIF data creates models with comparable snail distribution predictions and model performance measures as models given an expert-collected dataset. This is a very encouraging finding given that expert-executed field collection programs can be logistically infeasible and public species presence resources have grown in size and popularity (16). Moreover, even when expert field collection is feasible, it is often not possible to execute surveillance programs across large areas, such as the entirety of Brazil. GBIF cannot always guarantee the same level of species identification accuracy as the morphological and molecular tools often used in expert-executed field sampling, but the accessibility of large amounts of species data has dramatically increased the potential for species distribution analyses (16). Although only a singular case-study, our findings support the utility of GBIF data for producing accurate SDMs without targeted field collection programs. It is critical to employ methods to overcome spatial biases inherent in these publicly available data sources (42,43), such as through geographically stratified background sampling and careful inclusion criteria, but our findings support the growing use of these resources (26). Importantly, many of these conclusions rest on our example where there was sufficient quantity of GBIF data, which was only true of one species in São Paulo state. Our findings demonstrate the value that GBIF data can offer to disease control and elimination efforts and we support ongoing initiatives working to increase access, precision, and quality of GBIF data across all species and geographies (98).

Although our analysis contributes substantially to describing and quantifying current best practices in the SDM literature, there are several limitations. First, the smallest geographic extent we investigated was at the state level, which is still a large area. Other modeling studies, including some of specific Biomphalaria species, have been conducted at the municipality or intra-municipality scale (58,59). Although infeasible due to data quantity constraints across species for this study, it is possible that our comparisons could have been augmented for local specificity if we had included models built for specific municipalities. Secondly, we included a limited number of predictors in this case study for ease of interpretation, especially given our plan to compare models across geographic extents, machine learning models, and data sources. However, some of our findings could be sensitive to the number, resolution, and/or spatial-autocorrelation of predictors included (99). For example, a set of predictors dominated by LULC variables—rather than our models that included only two—could come to differing conclusions on changes in variable importance or partial dependence relationships. However, our set of predictors was chosen to be biologically relevant, sufficient to capture ecological relationships, and sufficiently general to be representative for other species distribution modeling studies. A combination of bioclimatic, LULC, and other variables is very common in the body of literature informing this analysis (33). Lastly, while this analysis does compare results across three species of snails, the species are very similar in that they are all from the same genus and are all freshwater mollusks. Other species, even among those relevant to disease ecology, could vary in their response to our analyses across machine learning models, spatial extents, and data sources (12). However, our analysis shows that even species in the same genus may have significantly different ecological niches, indicating that modeling decisions need to be grounded in system-specific ecological and biological knowledge.

There rightfully remains no single gold-standard of SDM methods suitable for all species, geographic locations, and applications because differing contexts and intended uses warrant differing modeling decisions. Making species distribution models that are useful and accurate for a given question of interest requires careful design and in-depth evaluation. This manuscript aims to serve as a resource and reference for current methods in species distribution modeling, with applications to disease ecology. Given the extent to which these models are used to inform fieldwork, policy, funding, and intervention strategies, continuous assessment and model evaluation are imperative. Species distribution models are powerful tools if used appropriately, and this work illustrates the importance of three key dimensions of variation—model type, spatial extent, and data source—highlighting that the former two can have large implications for model predictions and interpretation.

## Data Availability

All data and code used to construct the models described in this manuscript will be made publicly available upon publication.

## Acknowledgements

We would like to thank the Medical Malacology Collection of Fiocruz Minas for sharing *Biomphalaria* data used in this study.

## Supporting information captions

<u>Supplementary Table S1:</u> *B. straminea* presence data quantity by scale, thinned to 1 km.

<u>Supplementary Table S2:</u> Mean spatially cross-validated out-of-sample AUC, sensitivity, and specificity values for national models across machine learning model type. The corresponding table for in-sample estimates can be found in **Supplementary Table 3**.

<u>Supplementary Table S3:</u> In-sample AUC, sensitivity, and specificity values for national models of all species across machine learning model types.

<u>Supplementary Table S4:</u> In-sample AUC, sensitivity, specificity, and TSS measures calculated on state-level presence and background records for nationally-fit and state-specific models. *B. glabrata* and *B. straminea* were tested in Minas Gerais state, and *B. tenagophila* was tested in São Paulo state.

<u>Supplementary Figure S1:</u> Maps of environmental covariates across geographic scales (national, Minas Gerais, and São Paulo state). Bioclimatic variables (i.e. temperature and precipitation) are colored in orange, soil-related variables in blue, and land-use/land-cover variables in green.

A. Temperature seasonality (standard deviation of the monthly mean temperatures)
B. Mean daily temperature of the coldest quarter
C. Mean monthly precipitation of the wettest quarter
D. Mean monthly precipitation amount of the driest quarter
E. Soil clay percentage
F. Height above nearest drainage
G. Soil pH
H. Soil water percentage
I. Proportion of temporary crop cover
J. Distance to high population density

<u>Supplementary Figure S2:</u> *B. straminea* presence points by source (shape), thinned to 1 km. A) National, B) Minas Gerais, C) São Paulo

<u>Supplementary Figure S3:</u> National prediction maps of *B. straminea* (A–C) suitability probabilities by model type when models were provided the full set of species presence records available.

<u>Supplementary Figure S4:</u> Plots of ten-fold spatially cross-validated, out-of-sample AUC, TSS, sensitivity, and specificity values across all three species (A, B, C), scales (panels), and model types (colors). Plots display mean (point) and standard error (error bar).

<u>Supplementary Figure S5:</u> São Paulo prediction maps of *B. tenagophila* suitability probabilities by model type (rows) and model geographic extent (columns) when models were provided the full set of species presence records available.

<u>Supplementary Figure S6:</u> Minas Gerais prediction maps of *B. straminea* suitability probabilities by model type (rows) and model geographic extent (columns) when models were provided the full set of species presence records available.

<u>Supplementary Figure S7:</u> Proportion of total variable importance averaged across all training folds attributable to distance to high population density and proportion of temporary crop cover for *B. straminea*.

<u>Supplementary Figure S8:</u> Same as Figure 6 except all models are provided the same number of data points (*n* = 115, i.e. the number of total GBIF points for *B. tenagophila* in São Paulo). Data quantity was reduced through random selection when necessary. Spatially cross-validated AUC values for *B. tenagophila* São Paulo models (all reduced to *n* = 115): expert-collected mean AUC = 0.77, 10th/90th percentiles: [0.66, 0.91], publicly-available GBIF mean AUC = 0.79, [0.66, 0.93], and combined mean AUC = 0.75, [0.59, 0.96].

